# Favipiravir for treatment of outpatients with asymptomatic or uncomplicated COVID-19: a double-blind randomized, placebo-controlled, phase 2 trial

**DOI:** 10.1101/2021.11.22.21266690

**Authors:** Marisa Holubar, Aruna Subramanian, Natasha Purington, Haley Hedlin, Bryan Bunning, Katharine S. Walter, Hector Bonilla, Athanasia Boumis, Michael Chen, Kimberly Clinton, Liisa Dewhurst, Carol Epstein, Prasanna Jagannathan, Richard H. Kaszynski, Lori Panu, Julie Parsonnet, Elizabeth L. Ponder, Orlando Quintero, Elizabeth Sefton, Upinder Singh, Luke Soberanis, Henry Truong, Jason R. Andrews, Manisha Desai, Chaitan Khosla, Yvonne Maldonado

## Abstract

**Background:** Favipiravir is an oral, RNA-dependent RNA polymerase inhibitor with *in vitro* activity against SARS-CoV2. Despite limited data, favipiravir is administered to patients with COVID-19 in several countries.

**Methods:** We conducted a phase 2 double-blind randomized controlled outpatient trial of favipiravir in asymptomatic or mildly symptomatic adults with a positive SARS-CoV2 RT-PCR within 72 hours of enrollment. Participants were randomized 1:1 to receive placebo or favipiravir (1800 mg BID Day 1, 800mg BID Days 2-10). The primary outcome was SARS-CoV-2 shedding cessation in a modified intention-to-treat (mITT) cohort of participants with positive enrollment RT-PCRs. Using SARS-CoV-2 deep sequencing, we assessed favipiravir’s impact on mutagenesis.

**Results:** From July 8, 2020 - March 23, 2021, we randomized 149 participants with 116 included in the mITT cohort. The participants’ mean age was 43 years (SD 12.5) and 57 (49%) were women. We found no difference in time to shedding cessation by treatment arm overall (HR 0.76 favoring placebo, 95% confidence interval [CI] 0.48 – 1.20) or in sub-group analyses (age, sex, high-risk comorbidities, seropositivity or symptom duration at enrollment). We observed no difference in time to symptom resolution (initial: HR 0.84, 95% CI 0.54 – 1.29; sustained: HR 0.87, 95% CI 0.52 – 1.45). We detected no difference in accumulation of transition mutations in the viral genome during treatment.

**Conclusions:** Our data do not support favipiravir use at commonly used doses in outpatients with uncomplicated COVID-19. Further research is needed to ascertain if higher doses of favipiravir are effective and safe for patients with COVID-19.

**Trial registration number:** NCT04346628

**Summary:** In this phase 2 double-blind randomized controlled outpatient trial of favipiravir in asymptomatic or uncomplicated patients with COVID-19, we found no difference in time to shedding cessation or time to symptom resolution by treatment arm.

## Introduction

Favipiravir is an oral, RNA-dependent RNA polymerase (RdRp) inhibitor with a wide spectrum of activity, including *in vitro* activity against SARS-CoV2. In its active form, favipiravir is incorporated into nascent viral RNA by error-prone viral RdRp and disrupts RNA synthesis directly by chain termination or accumulation of deleterious mutations in the SARS-COV-2 genome.[1] Since 2014, favipiravir has been used in Japan and China for patients with drug-resistant influenza and boasts an established, well-characterized safety profile, making it an attractive potential therapeutic option for COVID-19.

Early data from some open-label trials suggested that favipiravir improved clinical and/or virologic outcomes in patients with COVID-19. [2, 3] Despite limited data, favipiravir was approved for use in patients with COVID-19 in some countries. We evaluated favipiravir’s efficacy in reducing viral shedding duration and improving symptoms in outpatients with uncomplicated COVID-19.

## Methods

### Study Design

We conducted a Phase 2, double-blind, randomized, placebo-controlled phase 2 trial at Stanford Healthcare, California. Stanford University School of Medicine Panel on Human Subjects in Medical Research approved the study protocol. An independent Data and Safety Monitoring Board (DSMB) reviewed the study design, clinical trial progress, study integrity, and safety data including interim analysis.

### Participants

We enrolled asymptomatic or symptomatic adults without respiratory distress who had a positive SARS-CoV-2 RT-PCR collected within 72 hours of enrollment. We excluded individuals who required renal replacement therapy, had liver impairment, were immunocompromised or taking immunosuppressing medications, or were pregnant or breast-feeding.

Participants were randomized 1:1 to favipiravir or placebo using block, REDCap-implemented, randomization stratified by age (>=50 and <50 years old) and sex. [4, 5]

### Procedures

Participants received placebo or favipiravir at doses of 1800 mg BID on Day 1, then 800mg BID on days 2-10. Favipiravir and placebo tablets were identical in appearance to maintain blinding.

We followed participants for 28 days and performed a clinical assessment and collected oropharyngeal (OP) swabs and blood samples at each visit. Staff-collected OP specimens underwent a reverse-transcription polymerase chain reaction assay (RT-PCR) (Viroclinics Biosciences, Rotterdam, The Netherlands). Anti-SARSCoV-2 serology was performed using a virus plaque reduction neutralization assay (Viroclinics Biosciences, Rotterdam, The Netherlands).

Patients self-collected daily anterior nasal swabs on days 1-10, 14, 21, and 28 and submitted them directly for RT-PCR testing with an assay that targeted the viral nucleocapsid gene’s N1 and N3 regions (Quest Diagnostics, Secaucus, New Jersey).

Patients also completed electronic daily symptom surveys and recorded temperature and oxygen saturation using study-provided devices; all data was collected using REDCap Cloud version 1.6 (REDCap Cloud, Encinitas, California).

### Outcomes

We defined the primary outcome, SARS-CoV-2 shedding cessation, as the time from enrollment to the first of two consecutive negative nasal RT-PCRs. We defined time until initial resolution of symptoms as time from randomization until the first of two consecutive days without symptoms. We defined time until sustained symptom resolution similarly, with the additional condition that symptoms remain resolved throughout the remainder of the study. Decreased taste/smell, mild fatigue, and mild cough were recorded, but excluded as symptoms for this analysis.[6] We censored participants who did not meet the symptom endpoint on their last completed survey. Additional secondary outcomes included incidence of hospitalizations or emergency department visits during the study and adverse events graded for severity.[7]

### Sample qPCR testing and sequencing protocols

To test whether favipiravir was acting as a mutagen, one of its mechanisms of action [1], we deep sequenced SARS-CoV-2 from residual day 1, 5 and 10 participant nasal swabs using an Illumina MiSeq platform (Supplementary Methods).

### Statistical analysis

We assessed virologic outcomes in a modified intention-to-treat (mITT) cohort, which included all randomized participants whose first available nasal RT-PCR result on days 1-3 was positive. We assessed symptom outcomes in a symptomatic (smITT) cohort, which included all randomized participants who reported at least one symptom at enrollment that was not mild cough, mild fatigue, or decreased taste/smell. We assessed safety endpoints in the ITT cohort. All analyses adjusted for age group and sex. Unless otherwise noted, all tests were two-sided and conducted at an alpha level of 0.05. Analyses were performed in R version 4.0.2.[4, 5]

#### Primary analysis

We used a Cox proportional hazards model to compare time until shedding cessation between treatment arms. The final test was performed at the alpha = 0.04999 level of significance allowing for an interim analysis. We censored participants who did not meet the endpoint on the last positive PCR result date and verified the proportional hazards assumption by examining Schoenfeld residuals.

#### Secondary analyses

We used a Cox proportional hazards model to compare initial and sustained symptom resolution between arms and Fisher’s Exact test to compare proportions.

We evaluated change in Cycle Threshold (Ct) from Day 1 to Day 7 and from Day 1 to Day 10 by treatment arm using generalized linear mixed effects regression models (GLMM, Supplementary Methods).

#### Post-hoc and efficacy sensitivity analyses

We added a statistical interaction term between treatment and these baseline characteristics to the primary efficacy model to test for effect modification: seropositivity; high-risk status; symptom onset within 3, 5, and 7 days of enrollment; age group; sex. We classified participants as high risk if they met any of these criteria: age ≥ 65, BMI ≥ 35, chronic kidney disease, diabetes mellitus, or age ≥ 55 and with one of these comorbidities: cardiovascular disease, hypertension, or chronic respiratory disease.

Interaction terms were also added to the sustained symptom resolution model for high-risk status and symptom onset within 3 and 5 days of enrollment. We reported p-values from a Wald test corresponding to the interaction terms and within-subgroup hazard ratios.

#### Sample size determination

Assuming 1:1 randomization and a two-sided log rank test at alpha□=□0.04999 level of significance for the final analysis, we anticipated 79 shedding cessation events, which provided 80% power to detect a hazard ratio of 2.03. We additionally assumed median of 14 days to shedding cessation in the control arm and 7 days in the treatment arm, a 3-month accrual period, a 4-week follow-up period after randomization of the last patient, and 10% drop out in the control arm. This enabled an interim analysis conducted at alpha□=□0.00001 to assess overwhelming efficacy after 50% of participants completed 24□hours of follow-up. We estimated that the total sample size required to achieve 79 events was 120 (60 participants per arm).

At interim review, the DSMB recommended increasing the sample size with the goal of 120 participants in the mITT cohort.

### Variant identification

We used the nfcore/viralrecon v.2.3dev bioinformatic pipeline to perform variant calling and to generate consensus sequences from raw reads (Supplementary Methods).[8] We predicted that favipiravir would impact viral diversity by study day 5 and result in a higher rate of transition mutations. [1, 9]

To assess favipiravir’s impact on SARS-CoV-2 within-host diversity, we tested if the number of iSNVs, transitions, and/or either iSNVs and transitions standardized by the total number of bases sequenced in a sample differed between the treatment arms on day 5 using one-sided two-sample t-tests with the R package rstatix.[10] We fit independent linear models for the number of iSNVs, standardized number of iSNVs, number of transitions, and standardized number of transitions with study day and treatment group as predictor variables in the R package stats.[11]

We used a p-value threshold of 0.05 to identify predictors significantly associated with within-host viral diversity.

## Results

From July 8, 2020 through March 23, 2021, we screened 385 patients and randomized 149 patients who were included in the ITT cohort (74 placebo, 75 favipiravir; Figure 1). Of these, 116 participants were included in the mITT and 135 in the smITT cohorts; 112 participants were included in all 3 analytic cohorts (Supplementary Figure 1).

**Figure 1.**
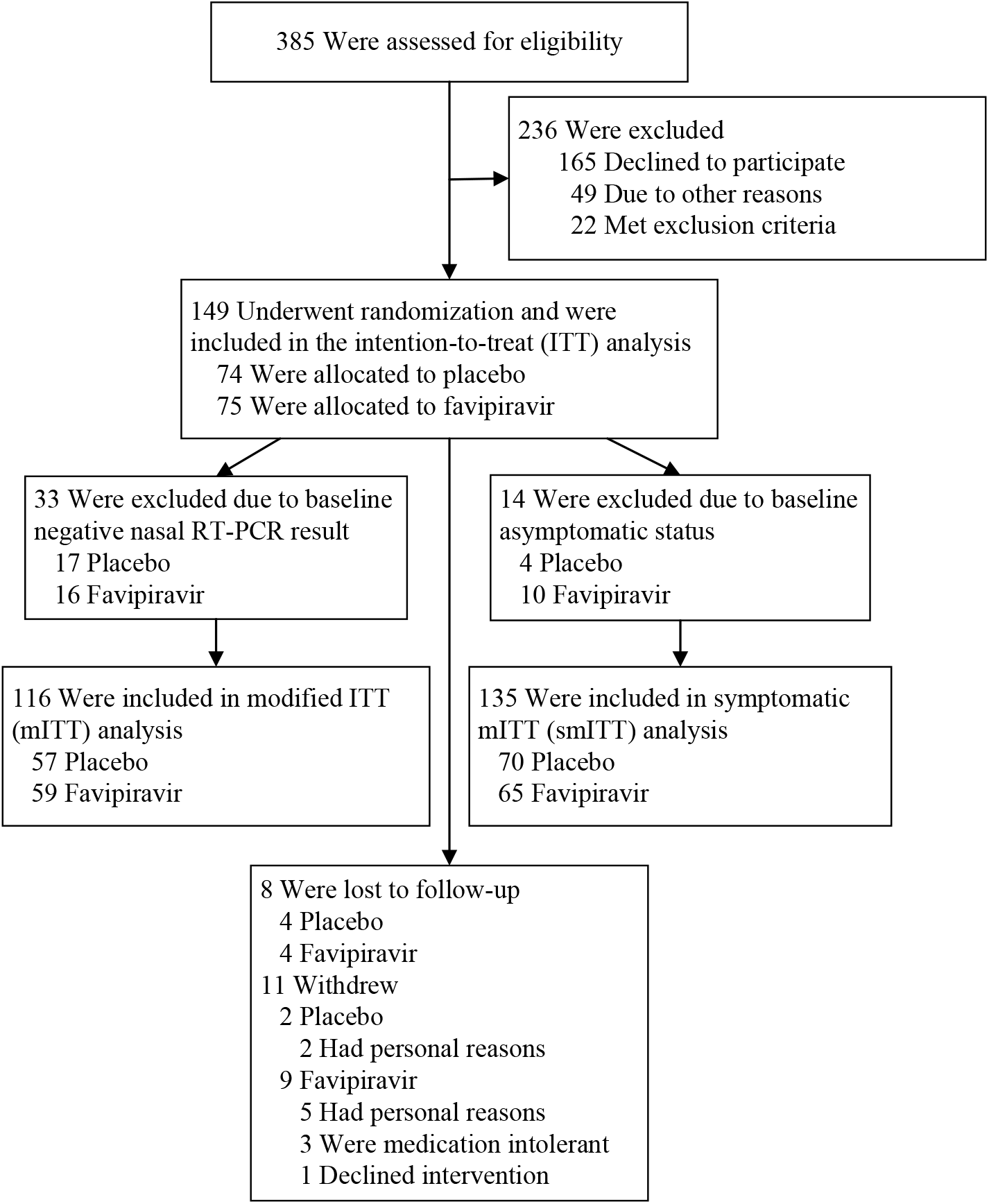
CONSORT diagram. Trial schematic showing participants screened, randomized, and followed through study completion.

Baseline demographic and disease characteristics were balanced between the two groups in all analytic cohorts (Table 1). In the mITT cohort, 31% of participants had at least one comorbidity of interest, and 37% had a body mass index >= 30. Of those with a positive RT-PCR upon enrollment, the median Ct was 24 [IQR 21-28] for the N1 target and only 10 participants had detectable antibodies (placebo 4, favipiravir 6).

**Table 1.** Baseline characteristics

|  | mITT (n=116) |  |  | smITT (n=135) |  |  |
| --- | --- | --- | --- | --- | --- | --- |
|  | Placebo<br>(n=57) | Favipiravir<br>(n=59) | SMD | Placebo<br>(n=70) | Favipiravir<br>(n=65) | SMD |
| **Age at randomization in years, mean (SD)** | 43.4 (12.8) | 42.9 (12.3) | 0.04 | 42.8 (12.6) | 42.5 (12.0) | 0.03 |
| **Female, n (%)** | 29 (50.9) | 28 (47.5) | 0.07 | 37 (52.9) | 32 (49.2) | 0.07 |
| **Race/ethnicity, n (%)** |  |  | 0.14 |  |  | 0.20 |
| Latinx | 24 (42.1) | 26 (44.1) |  | 29 (41.4) | 28 (43.1) |  |
| White | 21 (36.8) | 19 (32.2) |  | 26 (37.1) | 22 (33.8) |  |
| Asian | 5 (8.8) | 6 (10.2) |  | 7 (10.0) | 6 (9.2) |  |
| Native Hawaiian/<br>Pacific Islander | 1 (1.8) | 2 (3.4) |  | 1 (1.4) | 3 (4.6) |  |
| Other/Unknown | 6 (10.5) | 6 (10.2) |  | 7 (10.0) | 6 (9.2) |  |
| **Mean body mass index (BMI) (SD)** | 29.3 (6.0) | 27.8 (5.7) | 0.25 | 28.9 (5.9) | 28.0 (5.8) | 0.15 |
| **BMI 30+, n (%)** | 25 (43.9) | 18 (30.5) | 0.33 | 29 (41.4) | 21 (32.3) | 0.19 |
| **Number with comorbid conditions, n (%)** |  |  |  |  |  |  |
| None | 39 (68.4) | 41 (69.5) | 0.02 | 48 (68.6) | 47 (72.3) | 0.08 |
| Diabetes Mellitus | 3 (5.3) | 7 (11.9) | 0.24 | 4 (5.7) | 8 (12.3) | 0.23 |
| Hypertension | 5 (8.8) | 5 (8.5) | 0.01 | 8 (11.4) | 6 (9.2) | 0.07 |
| Chronic lung disease | 3 (5.3) | 2 (3.4) | 0.09 | 3 (4.3) | 2 (3.1) | 0.06 |
| **Asymptomatic, n (%)** | 1 (1.8) | 3 (5.1) | 0.18 | 0 | 0 | <0.01 |
| **Days from symptom onset to randomization, median [IQR]** | 5 [4, 6] | 5 [3, 7] | 0.01 | 5 [4, 7] | 5 [3, 7] | 0.08 |
| **Number of symptoms reported at randomization, median [IQR]** | 6 [4, 9] | 6 [4, 8.5] | 0.28 | 6 [4, 9] | 6 [4, 8] | 0.16 |
| **Symptoms at randomization, n (%)** |  |  |  |  |  |  |
| Fever | 2 (3.5) | 1 (1.7) | 0.11 | 3 (4.3) | 1 (1.5) | 0.16 |
| Cough/Dyspnea | 44 (77.2) | 42 (71.2) | 0.14 | 48 (68.6) | 47 (72.3) | 0.08 |
| Fatigue | 41 (71.9) | 40 (67.8) | 0.09 | 51 (72.9) | 47 (72.3) | 0.01 |
| Joint pain | 18 (31.6) | 20 (33.9) | 0.05 | 20 (28.6) | 22 (33.8) | 0.11 |
| Myalgias | 36 (63.2) | 36 (61.0) | 0.04 | 42 (60.0) | 38 (58.5) | 0.03 |
| Headache | 37 (64.9) | 40 (67.8) | 0.06 | 45 (64.3) | 43 (66.2) | 0.04 |
| **Received at least one dose of COVID-19 vaccine, n (%)** | 2 (3.5) | 0 (0.0) | 0.27 | 2 (2.9) | 0 (0.0) | 0.24 |
| **Baseline seropositivity, n (%)** | 4 (7.0) | 6 (10.2) | 0.30 | 11 (15.7) | 9 (13.8) | 0.14 |
| **Baseline anterior nares RT-PCR Ct, median, [IQR]** | 25.1<br>[22.2, 28.9] | 22.2<br>[19.7, 27.2] | 0.30 | 28.3<br>[23.2, 38.4] | 24.3<br>[20.7, 31.9] | 0.38 |
| **Baseline oropharyngeal RT-PCR positivity, n (%)** | 50 (87.7) | 54 (91.5) | 0.18 | 52 (74.3) | 53 (81.5) | 0.24 |
| **Baseline laboratory values, median [IQR]** |  |  |  |  |  |  |
| AST (units/L) | 32.0<br>[26.0, 42.5] | 29.0<br>[25.0, 34.0] | 0.39 | 29.5<br>[25.8, 39.3] | 29.0<br>[25.0, 34.0] | 0.31 |
| ALT (units/L) | 29.0<br>[20.0, 48.0] | 25.0<br>[19.5, 38.0] | 0.18 | 24.5<br>[18.8, 46.5] | 25.0<br>[19.0, 37.0] | 0.16 |
| Creatinine (mg/dL) | 0.8<br>[0.6, 1.0] | 0.8<br>[0.6, 1.0] | 0.09 | 0.8<br>[0.7, 1.0] | 0.8<br>[0.6, 1.0] | 0.12 |
| Uric acid (mg/dL) | 4.5<br>[3.5, 5.8] | 4.4<br>[3.9, 5.3] | <0.01 | 4.5<br>[3.5, 5.6] | 4.4<br>[3.9, 5.3] | 0.02 |

475 SMD = standardized mean difference; IQR = inner quartile range; Ct = cycle threshold

### Primary Analysis

Of the mITT population, 79 participants met the primary endpoint (44/57 [77%] placebo versus 35/59 [59%] favipiravir). Although the likelihood of shedding cessation favored placebo, we found no statistically significant difference in time to shedding cessation by treatment arm (HR 0.76, 95% confidence interval [CI] 0.48 – 1.20, P-value =0.24; Figure 2). We detected no difference in median time to shedding cessation between groups (placebo: 13 days (95% CI 9 – 14) versus favipiravir: 14 days (95% CI 9 – 21) Table 2). Of the 37 participants who did not meet the primary outcome, 18 had at least one negative RT-PCR during the study (8 placebo, 10 favipiravir).

**Table 2.** Primary and Secondary Outcomes

|  | Treatment arm |  | Measure of association |  |
| --- | --- | --- | --- | --- |
|  | Placebo | Favipiravir | aHR (95% CI) | p-value |
| **Primary Outcome<sup>2</sup>** |  |  |  |  |
| Days until viral shedding cessation, median (95% CI) | 13 (9, 14) | 14 (9, 21) | 0.76 (0.48, 1.20) | 0.24 |
| **Secondary Clinical Outcomes** |  |  |  |  |
| Hospitalizations by Day 28<sup>1</sup>, n participants (%) | 4/74 (5) | 0 | . | 0.06 |
| Emergency Department visits by Day 28<sup>1</sup>, n participants (%) | 7/74 (10) | 5/75 (7) | . | 0.56 |
| Days until initial resolution of symptoms<sup>3</sup>, median (95% CI) | 14 (11, 18) | 15 (12, 26) | 0.84 (0.54, 1.29) | 0.43 |
| Days until sustained resolution of symptoms<sup>3</sup>, median (95% CI) | 24 (21, NA) | NA (26, NA) | 0.87 (0.52, 1.45) | 0.59 |
| **Secondary Virologic Outcomes<sup>2</sup>** |  |  | **Δ inverse Ct (95% CI)** | **p-value** |
| Change in reverse Ct from Day 1 to 7, mean (SD) | -7.0 (5.6) | -9.2 (5.0) | -2.06 (-4.34, 0.22) | 0.08 |
| Change in reverse Ct from Day 1 to 10, mean (SD) | -10.5 (5.1) | -12.9 (5.9) | -1.83 (-4.19, 0.53) | 0.13 |
| Negative by RT-PCR on Day 7, n participants (%) | 10/47 (21) | 10/42 (24) | . | 0.80 |
| Negative by RT-PCR on Day 10, n participants (%) | 23/45 (51) | 20/35 (57) | . | 0.65 |
| **Safety Outcomes<sup>1</sup>** |  |  |  |  |
| Serious Adverse Events, n events (%) | 1 (1.4) | 0 | . |  |
| Resulting in death | 0 | 0 | . |  |
| Resulting in hospitalization | 1 (100.0) | 0 | . |  |
| Adverse Events, n events | 15 | 27 | . |  |
| Adverse Events, n participants (%) | 10 (13.5) | 19 (25.3) | . | 0.11 |
| Grade 3 Adverse Events, n (%) | 2 (13.3) | 2 (7.4) | . |  |
| Most common adverse events, n participants (%) |  |  |  |  |
| Dizziness | 2 (2.7) | 3 (4.0) | . |  |
| Nausea | 3 (4.1) | 1 (1.3) | . |  |
| Day 10 uric acid (mg/dL), median (IQR) | 4.9 (4.1, 6.0) | 7.4 (6.3, 9.0) | . |  |

<sup>1</sup> among the intention-to-treat (ITT) population.
<sup>2</sup> among the modified ITT population.
<sup>3</sup> among the symptomatic ITT population.
<sup>4</sup> among ITT population who were not seropositive at enrollment.
NA = undefined; aHR = adjusted hazard ratio (adjusted for age 50+ and sex); CI = confidence interval; Ct = cycle threshold; OP = oropharyngeal; RT-PCR = reverse transcription-polymerase chain reaction. All virologic endpoints use anterior nares swab results unless otherwise stated.

**Figure 2.**
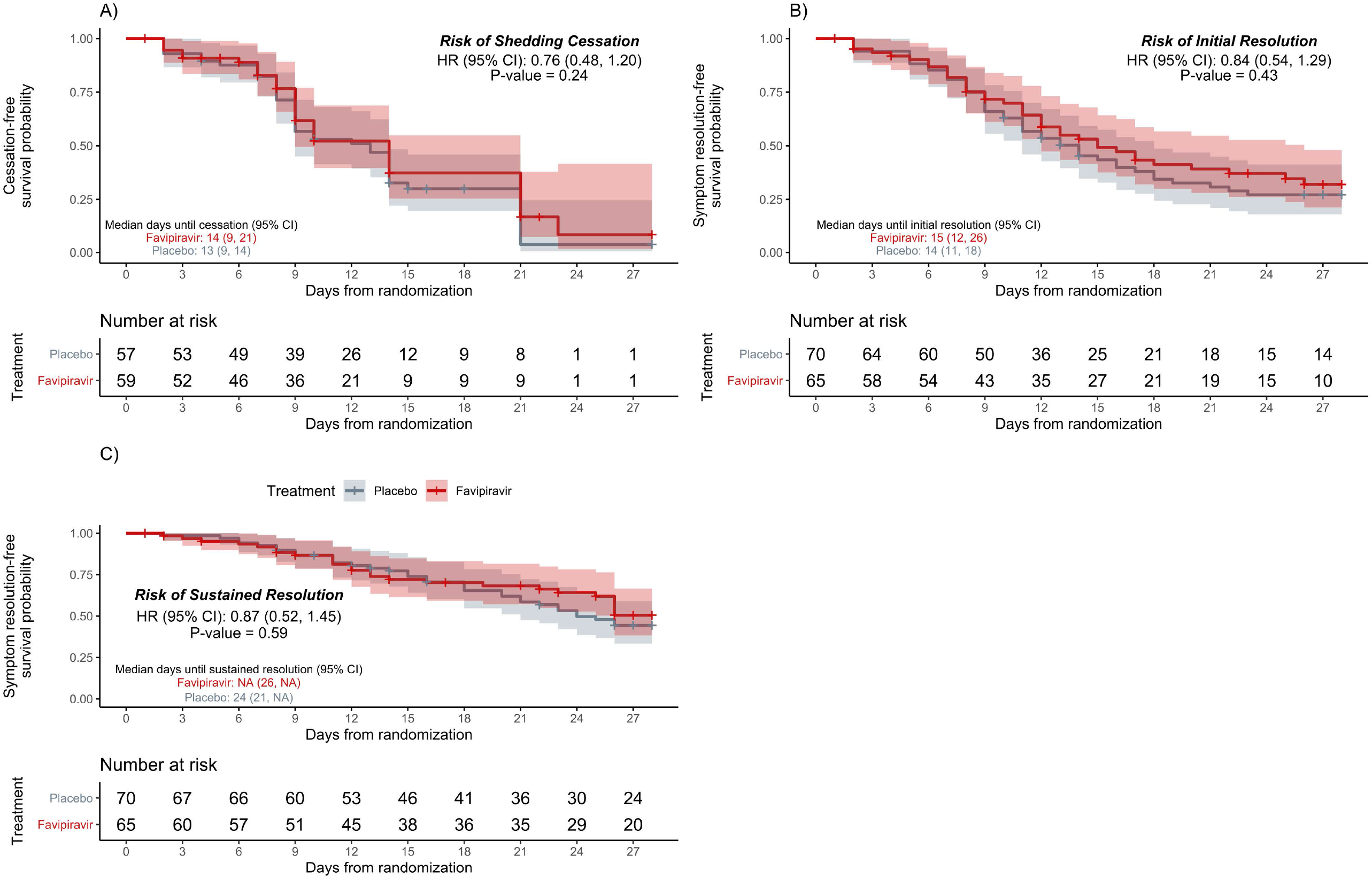
Kaplan–Meier analyses of the primary and key secondary outcomes in the modified intention-to-treat population. Time until a) shedding cessation of SARS-CoV-2 in RT-PCR from nasal swabs; b) initial symptom resolution; c) sustained symptom resolution stratified by treatment arm, favipiravir (red) vs. placebo (gray). Participants who did not experience the endpoint were censored (+ symbol) at their last positive swab for the primary outcome or at the last completed symptom questionnaire for the key secondary outcomes. Solid lines represent Kaplan–Meier survival probability; shading represents 95% confidence intervals.

In pre-specified and post-hoc analyses, we found no difference in time to shedding cessation by sub-groups including age group, sex, high risk comorbid conditions, seropositivity or duration of symptoms at enrollment (Supplementary Table 1).

In a sensitivity analysis using the ITT cohort, the median time to shedding cessation decreased to 9 days for both arms.

### Secondary Analyses

In the smITT population, both groups reported a median of 5 days of symptoms at enrollment (Table 1). The most common symptoms included cough/dyspnea, fatigue, myalgias, and headache.

We found no statistically significant difference in time to initial or sustained symptom resolution by treatment arm (initial: HR 0.84, 95% CI 0.54 – 1.29; sustained: HR 0.87, 95% CI 0.52 – 1.45; Table 2, Figure 2). The median time to initial symptom resolution was 1 day shorter in the placebo arm (14 days; 95% CI 11 – 18 versus 15 days; 95% CI 12 – 26). Although participants reported fewer and milder symptoms over time, 30 participants (18 placebo, 12 favipiravir) continued to report at least 1 symptom on day 28 (Figure 3, Supplementary Figures 3 and 4).

**Figure 3.**
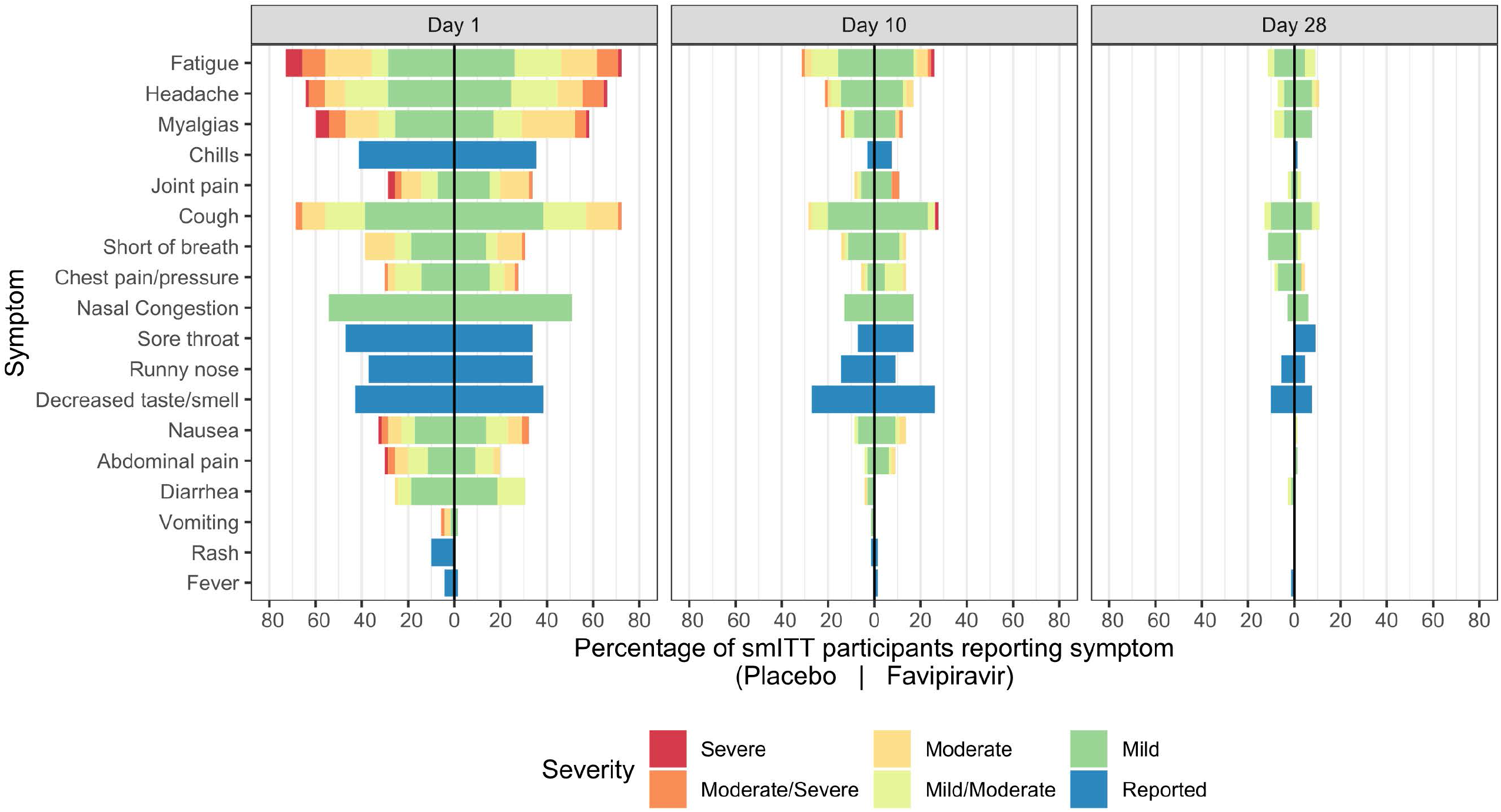
Symptom prevalence in the symptomatic modified intention-to-treat population. Mirrored bar plots of percentage of smITT participants reporting symptoms by treatment arm and study day, colored by symptom severity. Numerator is the number of participants reporting the symptom severity per study day and treatment arm; denominator is the number of overall participants in the treatment arm (n=70 in placebo and n=65 in favipiravir). Symptoms are ordered by Day 1 relative frequency within their respective organ systems (lower respiratory, upper respiratory, systemic, gastrointestinal, other). Bars to the right of the centered black line represent favipiravir symptom distributions, while those on the left are representative of placebo.

In the ITT cohort, 12 participants reported at least one emergency room visit during the study (7 (9.5%) placebo versus 5 (6.7%) favipiravir, p=0.56). Four were hospitalized and all 4 received placebo (Table 2).

Of the 124 randomized participants who did not have detectable antibodies at baseline, 71 (57%) were seropositive at day 28 (Supplementary Table 2).

### Virologic Analyses

Although the average Ct values increased significantly over time, the magnitude of decline did not differ between treatment arms (Figure 4, Supplementary Figure 2). We found no difference in the proportion of participants in either arm with a negative nasal RT-PCR on days 7 or 10 or a negative oropharyngeal RT-PCR on days 5 and 28 (Table 2, Supplementary Table 2).

**Figure 4.**
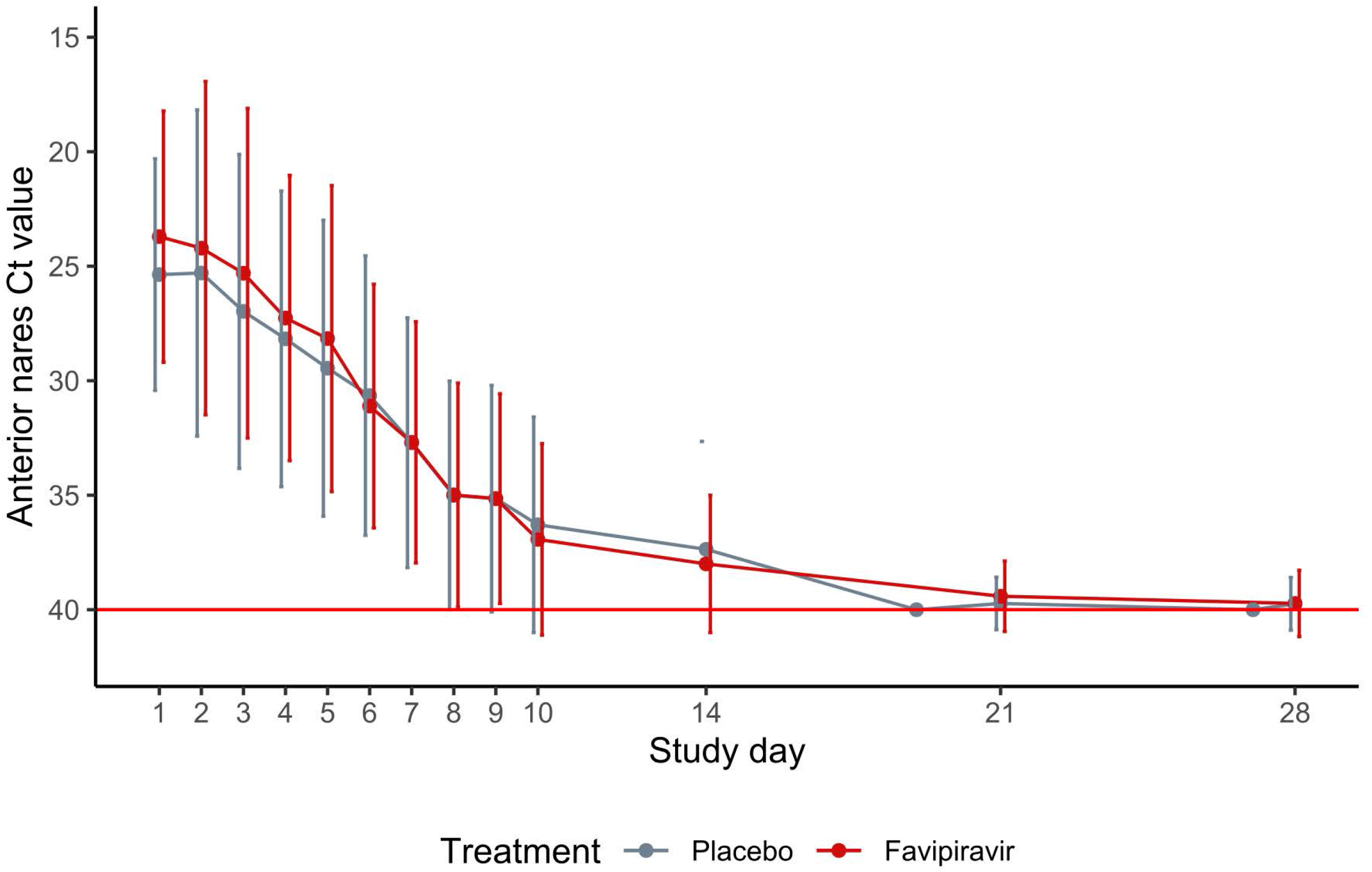
Trajectory of nasal cycle threshold in the modified intention-to-treat population. Line plots of nasal cycle threshold (Ct) values over time by treatment arm. Each dot represents the mean Ct value on that study day by treatment arm; bars represent the standard error around the mean. Lines are slightly jittered to avoid overlap. The red horizontal line at y=40 represents the limit of detection. Y-axis is reversed so that lower values of Ct represent more virus detected.

### Adverse Events

More participants reported adverse events in the favipiravir arm, but this difference was not statistically significant (10/71 (13.5%) in placebo vs. 19/75 (25.3%) in favipiravir arm; p=0.11; Table 2). The most common adverse event reported by those who received favipiravir was dizziness. More participants in the favipiravir arm developed hyperuricemia on study day 10 (placebo 21/71; 30% versus favipiravir 54/66; 82%) but only 3 participants were symptomatic.

### Sequencing Analyses

We included 112 PCR-positive nasal samples from 73 study participants (36 placebo, 37 favipiravir) that met our quality and coverage filters, including >1 longitudinal sample from 36 participants (18 placebo, 18 favipiravir). Residual nasal swabs had a mean qPCR CT of 22.3 and a mean depth of coverage of 1738X (95.1% of the genome with depth of coverage >10X). SARS-CoV-2 variation observed within a representative participant is shown in Supplementary Figure 5.

On day 5, we found no difference in the mean low frequency intrahost single nucleotide variants (iSNVs) in either arm (favipiravir 26.7 (std 16.5) versus placebo 37.4 (std 32.6), p= 0.23, two-sided t-test; Supplementary Figure 6). After standardizing by sequencing effort, the mean number of iSNVs was higher in the favipiravir (mean: 3.09 ×10^−8^ iSNVs/sequenced base-pairs; std: 3.24×10^−8^) compared to the placebo arm (mean: 2.1 ×10^−8^ iSNVs/bp; std: 2.03 ×10^−8^), but this difference was not significant (p = 0.35, two-sided t-test).

We found no difference in the number of transition iSNVs (p= 0.28, two-sided t-test) or the number of transition iSNVs standardized by sequencing effort (p= 0.37, two-sided t-test) in those who received favipiravir compared to placebo.

Finally, in linear models, we did not find that treatment arm was significantly associated with within-host SARS-CoV-2 diversity as measured by the raw number of iSNVs, the number of transition iSNVs, or the number of raw or transition iSNVs standardized by sequencing throughput, after controlling for study day.

## Discussion

In this trial of outpatients with asymptomatic or mild COVID-19, we found no difference in time to shedding cessation or symptom resolution between the favipiravir and placebo group.

Our results differ from previous open-label studies, possibly due to the added rigor of blinding and the robust data collection in our study. In an open-label favipiravir trial in India, Udwadia et al found no difference in time to viral shedding cessation using both oropharyngeal and nasopharyngeal swabs, however they did report a difference in time to clinical cure based on un-blinded clinician assessments of fever, oxygen saturation, and cough.[2] Our clinical symptom evaluation was more rigorous involving daily surveys which included a broader range of COVID-19 symptoms. In an open-label randomized controlled trial, Doi et al compared early (day 1) and late (day 6) favipiravir initiation and found a difference in fever resolution by day 2, but no difference in time to fever resolution or viral shedding.[12] In another open-label randomized controlled trial, Ivashenko et al found a difference in viral clearance by day 5 when they compared two favipiravir dosing regimens to standard of care, but this became equivalent by day 10.[3] Although we used a different primary outcome of time to shedding cessation as defined by 2 negative nasal RT-PCR tests, we also observed no difference in changes in RT-PCR Ct from day 1 to days 5 and 7.

To ensure robust outcomes, our study targeted those who were most likely to benefit from antiviral therapy by enrolling patients early in their illness. Overall, the median time from symptom onset to randomization was only 5 days, and in our mITT cohort only 10 out of 116 participants had developed anti-spike IgG at enrollment. Despite early favipiravir administration, we found no difference in either virologic or clinical outcomes.

We used the same favipiravir dosing regimen as other trials investigating favipiravir for COVID-19. [2, 3] In fact, some trials used the lower dosing regimen that is approved for patients with pandemic influenza in Japan. [13, 14] However, it is possible that this regimen did not achieve adequate levels to inhibit viral replication. A recent dose-optimizing study of 19 critically ill patients with influenza demonstrated a decrease in plasma trough concentrations (C_trough_) during the treatment course, estimating that only 42% of patients who received favipiravir 1800mg BID followed by 800 mg BID achieved the goal C_trough_ of ≥20 mg/L for >80% of the treatment duration.[15] Modeling from this work suggested that regimens of ≥3600 mg loading dose followed by 2600 mg might be necessary to achieve target concentrations. Trials investigating favipiravir for the treatment of Ebola used higher doses of favipiravir (6000mg/day load, then 2400mg/day), but also achieved lower drug concentrations than predicted at days 2 and 4 of treatment and did not meet their clinical endpoint.[16]

Suboptimal dosing may also explain why we found no evidence of mutagenesis after at least 5 days of favipiravir exposure. Our findings differ from *in vitro* work demonstrating a three-fold increase in the total number of mutations and twelve-fold increase in C to T or G to A transitions in Vero cells infected with SARS-CoV-2 exposed to favipiravir compared to controls.[1] This is also in contrast to a recent *in vivo* study of molnupiravir, a closely related nucleotide analogue, that found a two-fold increase in mutations in the SARS-COV-2 RdRp gene in the treatment compared to the control group.[17] A study that evaluated favipiravir dosing for Ebola infections in macaques found that viral mutational load was strongly associated with favipiravir dose[9] and that the accumulation of viral mutations was associated with lower levels of plasma infectious viral particles. Based upon these findings, the authors suggested that an earlier clinical trial in humans may have used suboptimal favipiravir dosing.

In contrast to our findings, an ongoing randomized placebo-controlled trial of molnupiravir —a nucleoside analog-prodrug--has reported a 50% reduction in COVID-19 related hospitalizations.[18] The overall hospitalization rates were higher than in our favipiravir study, possibly due to differences in standards of care and the predominance of SARS-CoV-2 B.1.617 (Delta variant) during the molnupiravir study. Of note, *in vitro* data suggests molnupiravir may also be mutagenic to mammalian cells.[19] Animal studies suggest that favipiravir administered in combination with molnupiravir may be an effective strategy to allow for lower molnupiravir doses and potentially avoid unintended consequences.[20]

Our study has several limitations. Most therapeutic studies for COVID-19, like ours, assess antiviral efficacy by using RT-PCR to detect viral RNA from nasal, nasopharyngeal or oropharyngeal swabs. However, detectable RNA may not reflect actively replicating virus and individuals can continue to have detectable RNA intermittently and long after illness recovery.[21] Widespread use of cell culture to detect replication-competent virus and to establish viral clearance is limited by feasibility, cost, and safety considerations.[21] Although we use cycle threshold rather than viral load, our analysis was strengthened by serial testing from individuals. Our primary endpoint was based upon participant-collected nasal swabs, which may be less accurate than nasopharyngeal swabs.[22] However, we found similar results from a secondary analysis of study staff collected oropharyngeal swabs. Our study was powered to detect differences in shedding cessation, not symptom resolution. Our study was not designed to detect a difference in long COVID syndrome, but we found that nearly half of both favipiravir and placebo-treated patients continued to report symptoms 28-day after enrollment. Finally, our study enrolled patients prior to the emergence and dominance of SARS-CoV-2 B.1.617 (Delta variant) in the US.

In conclusion, our data do not support favipiravir use at currently recommended doses in outpatients with mild or asymptomatic COVID-19. Dose optimization studies are necessary to elucidate if favipiravir administered at higher doses or delivered in combination with other agents is effective and safe for patients with COVID-19.

## Supporting information

FavipiravirSupplements

## Data Availability

All data produced in the present study are available upon reasonable request to the authors.

## Acknowledgements

We sincerely thank all participants for generously volunteering for this study and all study staff for their tireless effort and enthusiasm, including Renu Verma and Eugene Kim. We also thank our partners in Stanford Healthcare, Stanford University Occupational Health and San Mateo Medical Center in assisting in recruitment, Stanford University leadership for providing research space, and the DSMB members for their service. The study was funded by anonymous donors to Stanford University.

The Stanford REDCap platform (http://redcap.stanford.edu) is developed and operated by Stanford Medicine Research IT team. The REDCap platform services at Stanford are subsidized by a) Stanford School of Medicine Research Office, and b) the National Center for Research Resources and the National Center for Advancing Translational Sciences, National Institutes of Health, through grant UL1 TR001085. The Quantitative Sciences Unit is partially supported by UL1 TR003142.

## Funding

This work was supported by anonymous donors to Stanford University. The funders had no role in data collection, analysis, or the decision to publish.

## Notes

### Competing Interest Statement

The authors have declared no competing interest.

### Clinical Trial

NCT04346628

### Funding Statement

This study was funded by anonymous donors to Stanford University. The funders had no role in data collection, analysis, or the decision to publish.

### Author Declarations

IRB of Stanford University School of Medicine gave ethical approval for this work.

