## Supplementary material for "Favipiravir for treatment of outpatients with asymptomatic or uncomplicated COVID-19: a double-blind randomized, placebo-controlled, phase 2 trial": FavipiravirSupplements

| Table of Contents | Page number(s) |
|---|---|
| **Supplementary Methods** | **2** |
| **Supplementary Tables** | |
| Supplementary Table 1. Effect Modification Results | **4** |
| Supplementary Table 2. Additional Exploratory and Post-Hoc Results | **5** |
| **Supplementary Figures** | |
| Supplementary Figure 1. Venn Diagram of Cohorts | **6** |
| Supplementary Figure 2. Change in Ct from Day 1 to Days 7 and 10 (mITT) – box plot | **7** |
| Supplementary Figure 3. Participant-Level Summary of Symptoms by Initial Symptom Resolution | **8** |
| Supplementary Figure 4. Participant-Level Summary of Symptoms by Sustained Symptom Resolution | **10** |
| Supplementary Figure 5. SARS-CoV-2 Variation within a Study Participant | **11** |
| Supplementary Figure 6. Longitudinal trends in within-host SARS-CoV-2 diversity | **12** |

### Supplementary Methods

#### Change in Cycle Threshold over time

We evaluated change in Cycle Threshold (Ct) from Day 1 to Day 7 and from Day 1 to Day 10 by treatment arm using generalized linear mixed effects regression models (GLMM). Reverse Ct was defined by subtracting the Ct value from 40 (the limit of detection). Reverse Ct was fit as a function of treatment arm, categorical time (Day 7 or Day 10 versus Day 1) and the interaction between arm and time, adjusting for age group and sex with a random intercept to account for correlation within a participant. The interaction effects (along with corresponding 95% confidence intervals) between treatment arm and each time point were reported and reflected differences in change in reverse Ct by treatment arm. Using similar methodology (GLMM), reverse Ct was also modeled over time through Day 10 to assess whether the trajectory of reverse Ct differed by treatment arm. Unlike the former reverse Ct model, time was included as a continuous term and allowed for non-linear changes over time. More specifically, reverse Ct was fit to treatment arm, time, $time^2$, arm × time, arm × $time^2$, and adjusted for age and sex to account for randomization stratification variables. Model fit was assessed based on visual inspection of residuals.

#### Exploratory analyses.

We analyzed reverse SARS-CoV-2 Ct change over time through Day 10 using the OP PCR data and the same GLMM framework.

We used the Pearson chi-square test to compare the proportion of participants who developed SARS CoV-2 antibodies among those who were seronegative at enrollment between arms.

#### Sensitivity analyses

In a sensitivity analysis we compared shedding cessation using RT-PCR data from staff-collected OP samples to that from participant collected nasal swabs (used in primary endpoint). The proportion of participants who were RT-PCR positive among the mITT participants at Day 5 and Day 10 were compared across arms using the Pearson Chi-square test or the Fisher's exact test when the assumptions of the Chi-square test were not met. We also evaluated change in Ct using data RT-PCR from OP samples using similar statistical methods.

#### Sample qPCR testing and sequencing protocols

We deep sequenced SARS-CoV-2 from residual nasal swabs obtained from patients on days 1, 5 and 10. We extracted RNA from residual nasal swabs using the MagMAX™ Ultra Kit (Applied Biosystems), followed the CDC real-time qPCR N-gene assay to detect and quantify SARS-CoV-2 and quantified human RNaseP for quality control.[1] We followed the ARTIC v3 Illumina library preparation and sequencing protocols[2] and sequenced amplicons on an Illumina MiSeq platform.

#### Variant identification

Briefly, we used the nfcore/viralrecon v.2.3dev pipeline to remove reads mapping to the host genome with Kraken2,[3] align reads to the MN908947.3 Wuhan-1 reference genome with Bowtie 2[4], remove primer sequences with iVar[5], call variants with respect to the reference genome with iVar[5], generate a consensus sequence with iVar[5], and assign Nextclade lineages.[6] We sequenced six samples in duplicate; we combined reads in these duplicated samples for analysis. We focused our analysis on intrahost single nucleotide variants (iSNVs) and excluded any iSNVs within previously reported problematic sites.[7] We included samples with a median coverage of 100X and with >70% of the genome covered by a depth of >10X. We included iSNVs at genomic positions that had a total depth >100X, with at least two reads mapping to both the reference and alternate allele.

**Supplementary Table 1. Effect Modification Results For Viral and Sustained Symptom Cessation Endpoints**

| | | Viral shedding cessation | | Sustained symptom cessation | |
|---|---|---|---|---|---|
| **Effect modifier** | **Subgroup** | **Interaction p-value** | **aHR (95% CI) for favipiravir vs placebo** | **Interaction p-value** | **aHR (95% CI) for favipiravir vs placebo** |
| Baseline seropositivity | Seropositive | 0·84 | 0·76 (0·45, 1·28) | ·· | ·· |
| | Seronegative | | 0·86 (0·31, 2·34) | | ·· |
| High risk† | High risk | 0·25 | 0·67 (0·40, 1·15) | 0·68 | 0·80 (0·40, 1·14) |
| | Not high risk | | 1·26 (0·51, 3·12) | | 1·07 (0·31, 3·70) |
| Symptom onset within 3 days prior to enrollment† | ≤ 3 days | 0·30 | 0·82 (0·48, 1·40) | 0·48 | 0·90 (0·48, 1·40) |
| | > 3 days/asymptomatic | | 0·47 (0·19, 1·14) | | 0·60 (0·23, 1·52) |
| Symptom onset within 5 days prior to enrollment† | ≤ 5 days | 0·07 | 1·28 (0·63, 2·59) | 0·49 | 1·08 (0·63, 2·59) |
| | > 5 days/asymptomatic | | 0·54 (0·29, 0·98) | | 0·74 (0·38, 1·46) |
| Symptom onset within 7 days prior to enrollment† | ≤ 7 days | 0·94 | 0·74 (0·23, 2·40) | ·· | ·· |
| | > 7 days/asymptomatic | | 0·78 (0·47, 1·28) | | ·· |
| Age Group | 50+ | 0·51 | 0·69 (0·40, 1·19) | ·· | ·· |
| | <50 | | 0·97 (0·42, 2·25) | | ·· |
| Sex | Male | 0·06 | 0·51 (0·27, 0·95) | ·· | ·· |
| | Female | | 1·22 (0·63, 2·36) | | ·· |

† post-hoc analysis.

aHR = adjusted hazard ratio (adjusted for age group and sex); CI = confidence interval.

**Supplementary Table 2. Additional Exploratory and Post-Hoc Results**

| | Treatment arm | | Measure of association | |
|---|---|---|---|---|
| | **Placebo** | **Favipiravir** | **aHR (95% CI)** | **p-value** |
| **Exploratory Outcome** | | | | |
| Developed antibodies by Day 28[1], n participants (%) | 37/60 (62) | 34/64 (53) | ·· | 0·44 |
| **Post-Hoc Outcomes[2]** | | | | |
| Days until OP viral shedding cessation, median (95% CI) | 10 (10, 14) | 10 (10, 14) | 0·87 (0·55, 1·39) | 0·56 |
| | | | **Δ reverse Ct (95% CI)** | **p-value** |
| Change in inverse OP Ct from Day 1 to 5, mean (SD) | -4·3 (6·0) | -4·7 (6·4) | -0·39 (-2·44, 1·65) | 0·71 |
| Change in inverse OP Ct from Day 1 to 10, mean (SD) | -8·9 (7·3) | -7·9 (6·1) | 0·78 (-1·29, 2·85) | 0·46 |
| Negative by OP RT-PCR on Day 5 | 18/54 (33) | 20/55 (36) | ·· | 0·90 |
| Negative by OP RT-PCR on Day 10 | 46/55 (84) | 30/50 (60) | ·· | <0·001 |
| Negative by OP RT-PCR on Day 28 | 41/50 (82) | 42/46 (91) | ·· | ·· |

[1] among the intention-to-treat (ITT) population who were not seropositive at enrollment.

[2] among the modified ITT population.

aHR = adjusted hazard ratio (adjusted for age 50+ and sex); CI = confidence interval; OP = oropharyngeal; Ct = cycle threshold; RT-PCR = reverse transcription-polymerase chain reaction.

**Supplementary Figure 1. Venn diagram of analytic cohorts**

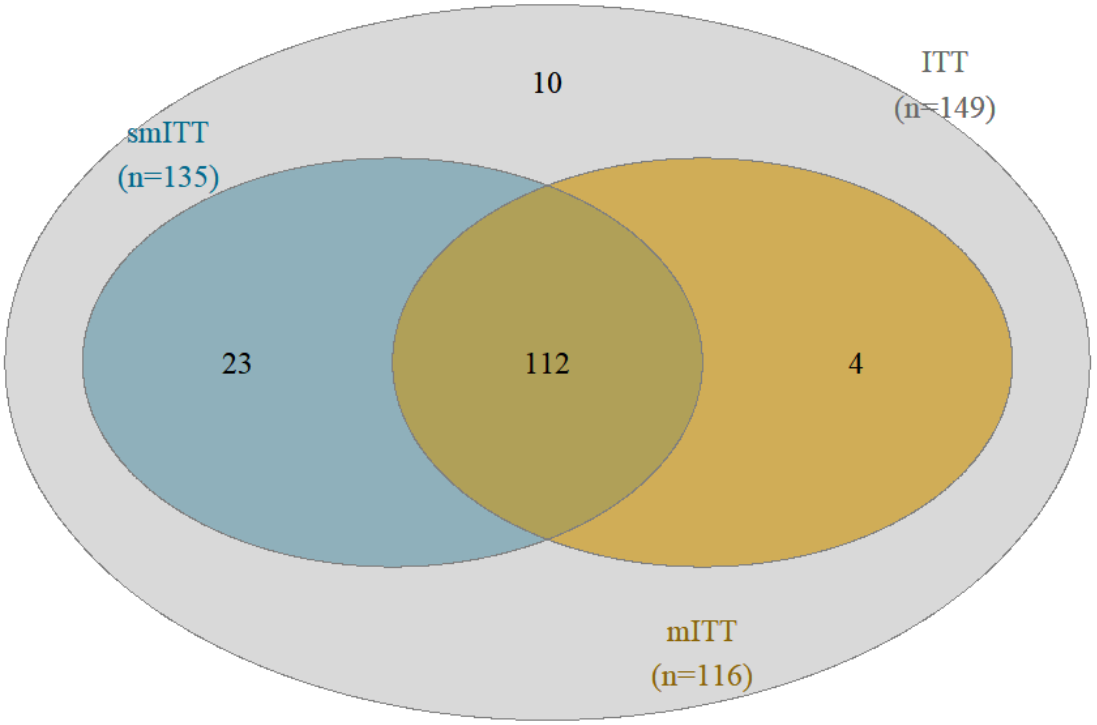

Caption: Venn diagram showing the overlap of participants among the intention-to-treat (ITT, gray), modified ITT (mITT, gold) and symptomatic ITT (smITT, blue) populations.

**Supplementary Figure 2. Change in Anterior Nares Reverse Cycle Threshold From Day 1 to Days 7 and 10 in the modified intention-to-treat population**

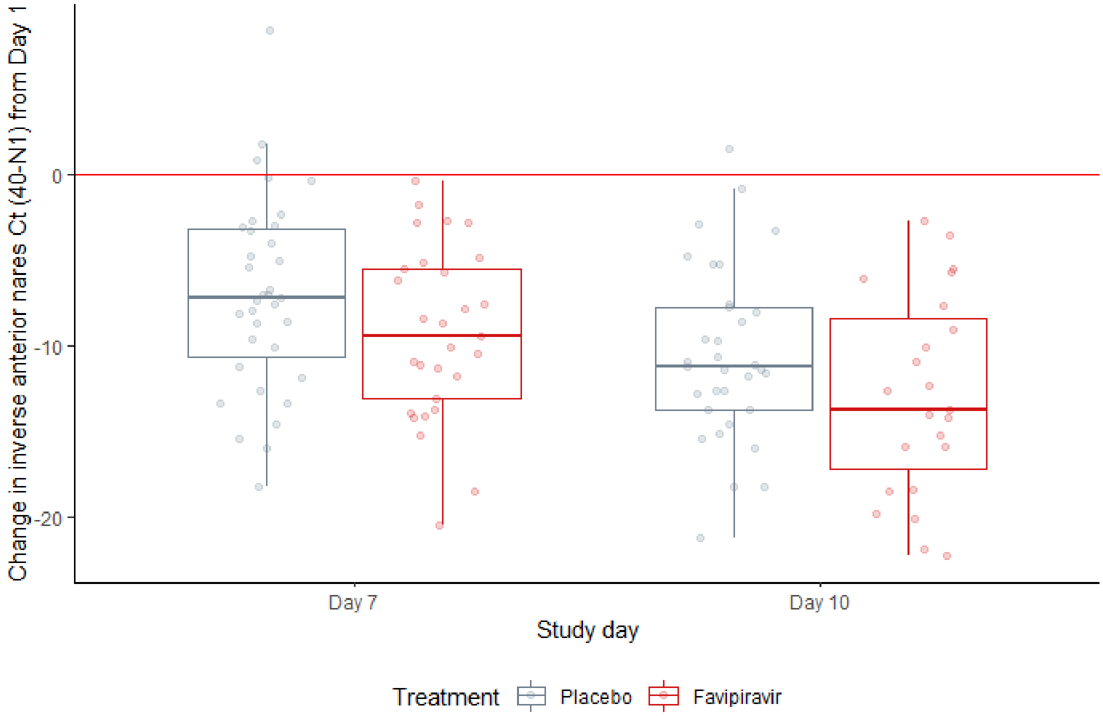

Caption: Boxplots of change in inverse anterior nares cycle threshold (Ct) from Day 1 to Days 7 and 10 by placebo (gray) and favipiravir (red). Dots represent participant-level values. The red reference line at zero represents no change from Day 1. Lower change values indicate lower viral load at subsequent study days.

**Supplementary Figure 3. Participant-Level Summary of Symptoms by Initial Symptom Resolution**

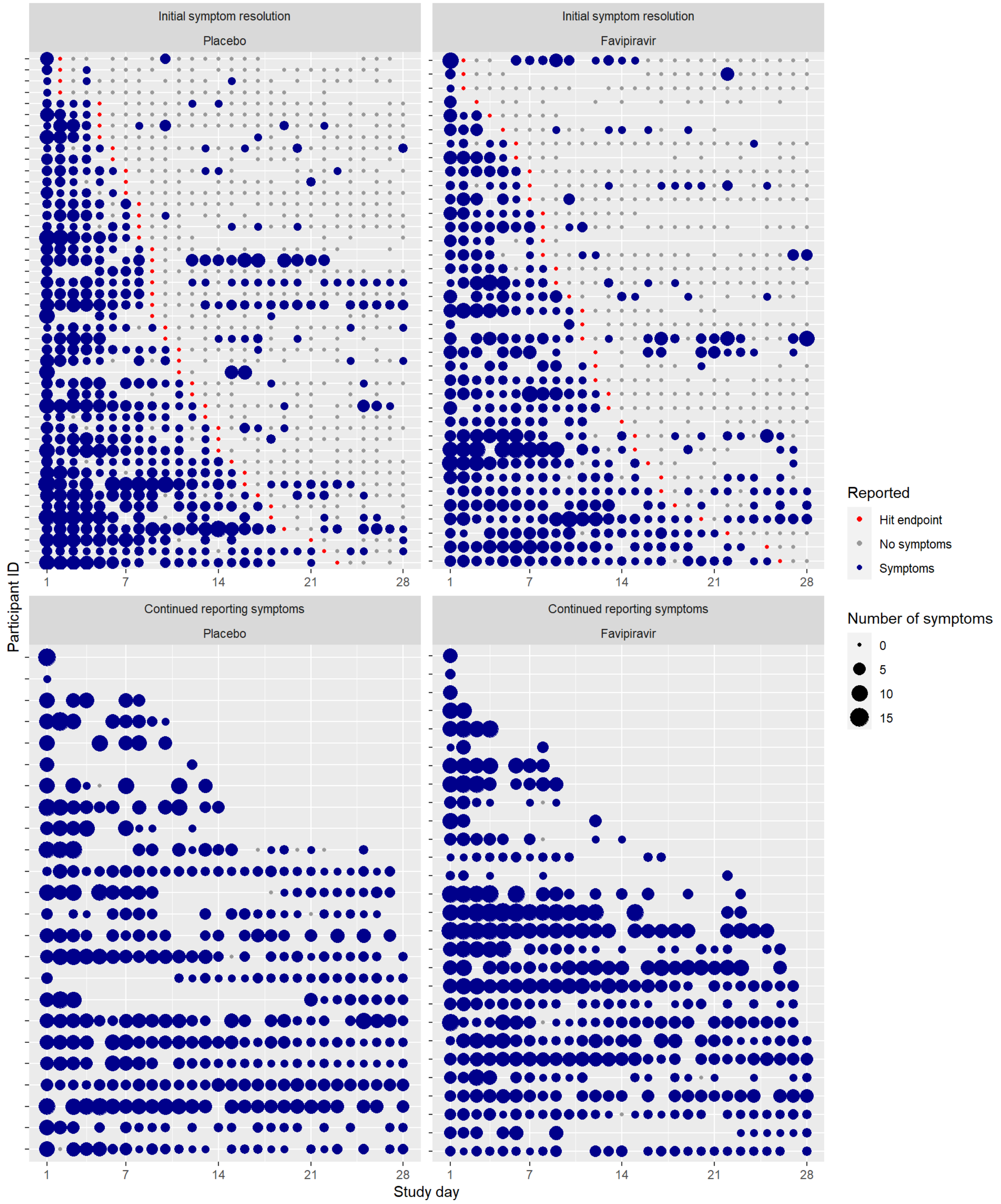

Caption: Participant-level dot plots of symptoms faceted by treatment arm and whether the initial symptom resolution endpoint was met. Dot size represents the number of symptoms reported on that study day while dot color represents whether symptoms were reported (blue), not reported (gray), and on which study day initial symptom resolution was met among those that met this endpoint (red). A missing data point indicates a symptom survey was not completed on that study day.

**Supplementary Figure 4. Participant-Level Summary of Symptoms by Sustained Symptom Resolution**

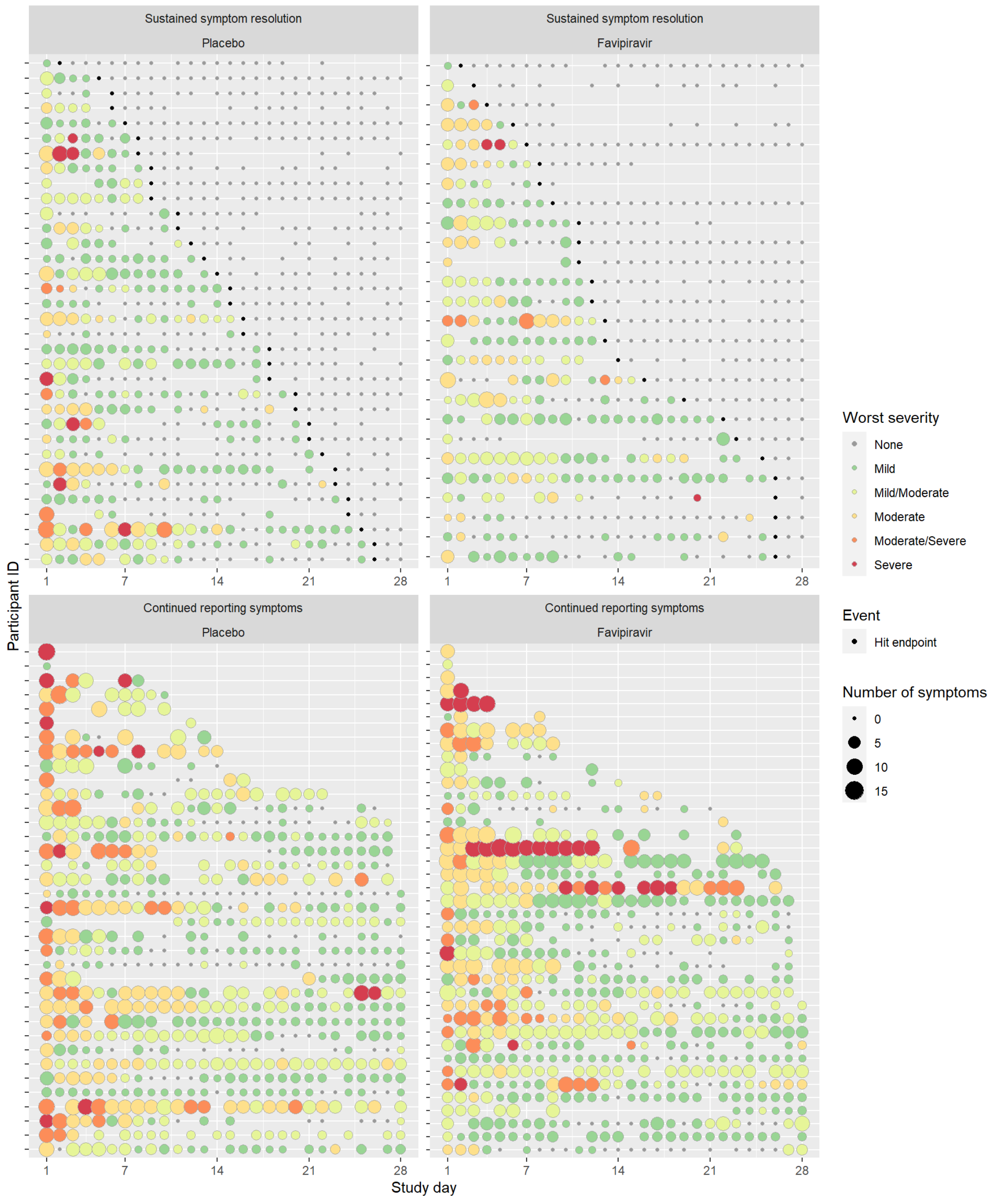

Caption: Participant-level dot plots of symptoms faceted by treatment arm and whether the sustained symptom resolution endpoint was met. Dot size represents the number of symptoms reported on that study day while dot color represents either the worst severity reported on that study day, or the day that the sustained symptom resolution endpoint was met among those who met this endpoint (black). A missing data point indicates a symptom survey was not completed on that study day.

**Supplementary Figure 5. SARS-CoV-2 Variation within a Study Participant.**

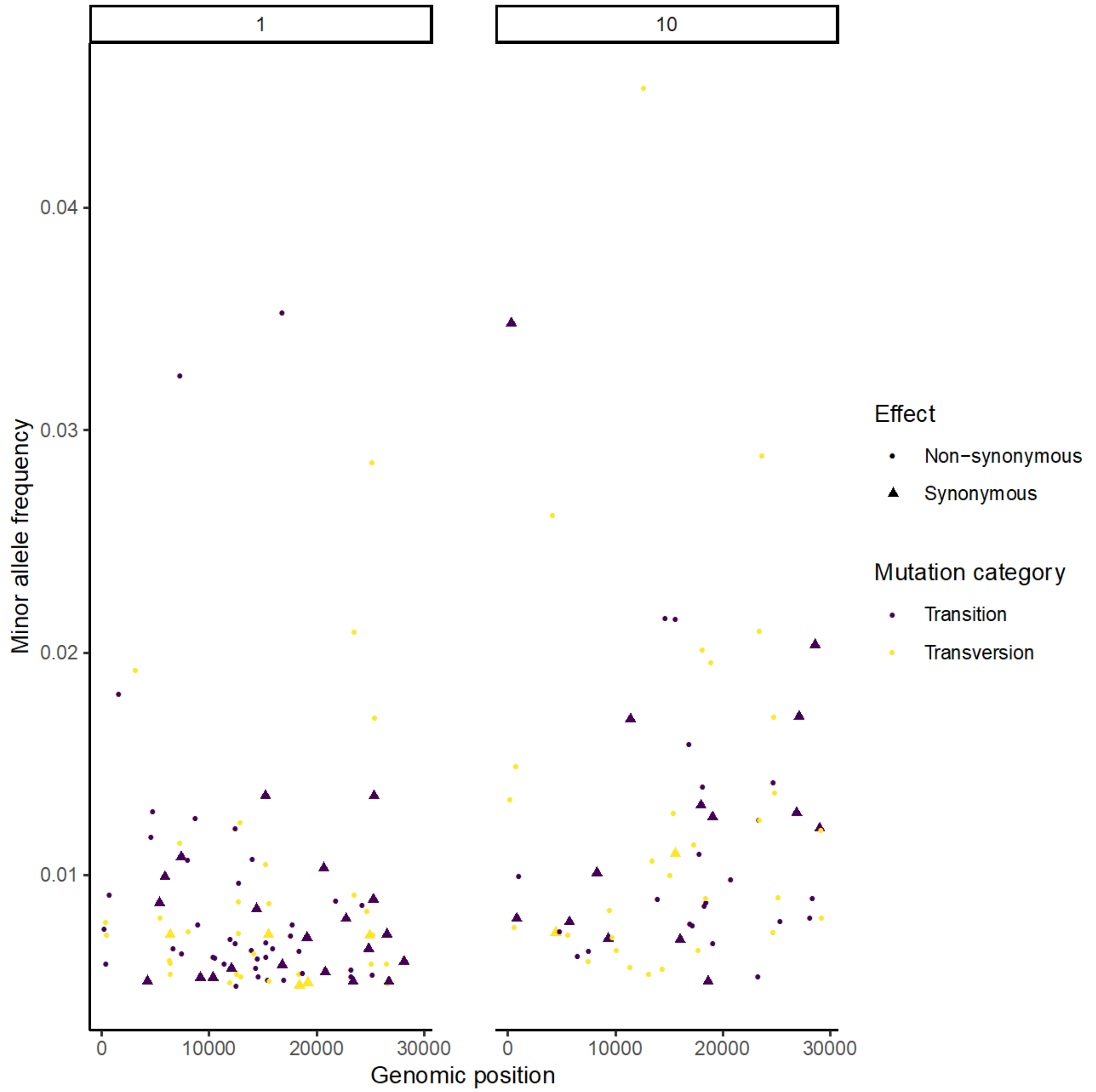

For a single representative participant in the Favipiravir arm, points indicate the minor allele frequencies of intrahost single nucleotide variants (iSNVs) identified along the SARS-COV-2 genome. Point shape indicates the predicted effect of each iSNV and color indicates the mutation category. Facets indicate study day.

**Supplementary Figure 6. Longitudinal Trends in within-host SARS-CoV-2 Diversity**.

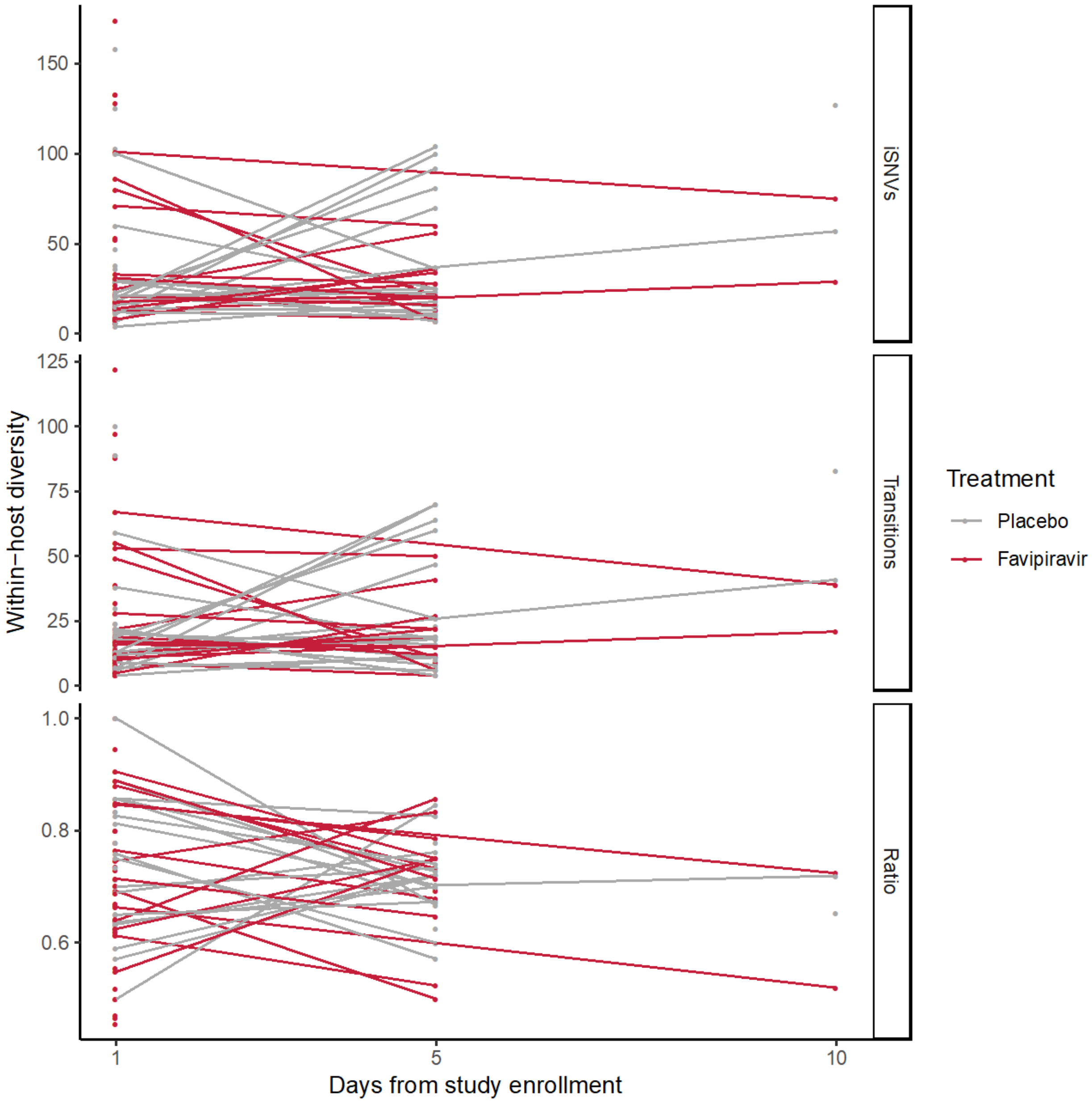

We quantified SARS-CoV-2 diversity in nasal swabs from study participants as the number of unique intrahost single nucleotide variants (iSNVs); the number of iSNVs that are transitions (Transitions); and the proportion of transitions among all iSNVs (Ratio). Lines represent unique participants sampled over time, points indicate unique samples and colors indicate study arm. We found no difference in the mean low frequency intrahost single nucleotide variants (iSNVs) in either treatment arm (favipiravir 26.7 (std 16.5) versus placebo 37.4 (std 32.6), p= 0.23, two-sided t-test.)
